# Post-COVID-19 symptoms are not uncommon among recovered patients-A cross-sectional online survey among the Indian population

**DOI:** 10.1101/2021.07.15.21260234

**Authors:** Guduru Venkat Rao, Vishwanath Gella, Madhuri Radhakrishna, Jagdeesh Kumar V, Robin Chatterjee, Anand V. Kulkarni, D. Nageshwar Reddy

**Author notes:** **Correspondence to:** Dr. Anand V. Kulkarni. MD, DM., Department of Gastroenterology and Hepatology, Asian Institute of Gastroenterology, Hyderabad, India. E-mail. **Financial disclosures:** None.

## Abstract

**Background:** Coronavirus disease-2019 (COVID-19) can have a myriad of symptoms. However, it is now known that most patients recovered from COVID-19 have symptoms related to COVID-19. There is a paucity of literature on post-COVID-19 symptoms from India. Hence we aimed to assess the incidence of post-COVID-19 symptoms in patients recovered from COVID-19. Methods: An online Microsoft forms survey was conducted through multiple social media platforms.

**Results:** Of the 5,347 individuals who received and clicked the link, a total of 2038 infected patients responded (Supplementary figure). Approximately 48% (967/2038) had recovered from COVID-19 within 1-3 months (short-term recovered), 34.2% (375/2038) had recovered from COVID-19 >3 months ago (long recovered), and 18.4% (375) were recovered within the last one month (recently recovered). Nearly 38% (770/2038) had a history of hospitalization for COVID-19. Of them, 34.28% (264/770) required oxygen therapy during the hospital stay. Most patients were discharged within 5-10 days of hospital stay (54%, 415/770). Only 5.58% (43/770) required a stay of more than 20 days. Seventy-five percent (575/770) of the hospitalized patients received steroid therapy. Of those who received steroid therapy, 56.5% (325/575) had not required oxygen therapy. Forty percent (233/575) of patients received steroid therapy for two weeks, 24% (138/575) for one week, 33.73% received steroids only during the hospital stay, and 1.73% were still on steroid therapy during the survey.

Most importantly, of the 2038 respondents, 41.8% (851/2038) still had persistent symptoms related to COVID-Most common symptom was fatigue (64.15%), followed by body pain (31%) and gastrointestinal symptoms (25%) (**Figure**). Six percent (49/851) of them required hospitalization for post-COVID-19 symptoms. Forty-six percent (449/967) in the short term recovered group (1-3 months), 40.1% (279/696) in the long-recovered group, and 32.8% (123/375) in the recently recovered group had persistent symptoms related to COVID-19 (P=0.001). Forty-eight percent (374/770) of the hospitalized patients developed post-COVID-19 symptoms, while only 37.6% (477/1268) developed post-COVID-19 symptoms among the non-hospitalized patients (P<0.001). Fifty-three percent (303/575) of those who received steroids developed post-COVID-19 symptoms, while only 36.41% (71/195) of those who did not receive steroids developed post-COVID-19 symptoms (P<0.001). 49% (159/325) of patients who received steroids despite being not requiring oxygen developed post-COVID-19 symptoms compared to only 37.5% (543/1449) who did not receive steroids and were not on oxygen therapy (P<0.001). Nearly 40% (336/851) of respondents felt that post-COVID-19 symptoms are not being appropriately treated or taken care of seriously.

**Conclusions:** Post-COVID-19 symptoms are common in patients who recovered from COVID-19. These symptoms are more often noted in patients who received steroid therapy. Post-COVID-19 symptomatology is present in a significant number of patients and requires to be addressed seriously.

## Introduction

Coronavirus disease-2019 (COVID-19) can have a myriad of symptoms. However, it is now known that most patients recovered from COVID-19 have symptoms related to COVID-19 ^1,2^. There is a paucity of literature on post-COVID-19 symptoms from India. Hence we aimed to assess the incidence of post-COVID-19 symptoms in patients recovered from COVID-19.

## Methods

An online Microsoft forms survey was conducted through multiple social media platforms. (Questionnaire in Appendix). The institutional ethics committee approved the study vide letter no. AIG/IEC-Post BH&R 15/06/2021-07. Categorical variables are expressed as n (%). The categorical data were compared between two groups using Pearson’s Chi-square test. A P-value <0.05 was considered significant. Logistic regression analysis was performed to assess the predictors of post-COVID-19 symptoms using a constant model.

## Results

Of the 5,347 individuals who received and clicked the link, a total of 2038 infected patients responded (**Supplementary figure**). Approximately 48% (967/2038) had recovered from COVID-19 within 1-3 months (short-term recovered), 34.2% (375/2038) had recovered from COVID-19 >3 months ago (long recovered), and 18.4% (375) were recovered within the last one month (recently recovered). Nearly 38% (770/2038) had a history of hospitalization for COVID-19. Of them, 34.28% (264/770) required oxygen therapy during the hospital stay. Most patients were discharged within 5-10 days of hospital stay (54%, 415/770). Only 5.58% (43/770) required a stay of more than 20 days. Seventy-five percent (575/770) of the hospitalized patients received steroid therapy. Of those who received steroid therapy, 56.5% (325/575) had not required oxygen therapy. Forty percent (233/575) of patients received steroid therapy for two weeks, 24% (138/575) for one week, 33.73% received steroids only during the hospital stay, and 1.73% were still on steroid therapy during the survey.

Most importantly, of the 2038 respondents, 41.8% (851/2038) still had persistent symptoms related to COVID-19. Most common symptom was fatigue (64.15%), followed by body pain (31%) and gastrointestinal symptoms (25%) (**Figure**). Six percent (49/851) of them required hospitalization for post-COVID-19 symptoms. Forty-six percent (449/967) in the short term recovered group (1-3 months), 40.1% (279/696) in the long-recovered group, and 32.8% (123/375) in the recently recovered group had persistent symptoms related to COVID-19 (P=0.001). Forty-eight percent (374/770) of the hospitalized patients developed post-COVID-19 symptoms, while only 37.6% (477/1268) developed post-COVID-19 symptoms among the non-hospitalized patients (P<0.001). Fifty-three percent (303/575) of those who received steroids developed post-COVID-19 symptoms, while only 36.41% (71/195) of those who did not receive steroids developed post-COVID-19 symptoms (P<0.001). 49% (159/325) of patients who received steroids despite being not requiring oxygen developed post-COVID-19 symptoms compared to only 37.5% (543/1449) who did not receive steroids and were not on oxygen therapy (P<0.001). Nearly 40% (336/851) of respondents felt that post-COVID-19 symptoms are not being appropriately treated or taken care of seriously. On univariate logistic regression analysis, hospitalized patients had higher odds of developing post-COVID-19 symptoms (OR,1.56 [1.3-1.87];P<0.001). Those who required oxygen therapy had a high chance of developing post-COVID-19 symptoms (1.97 [1.52-2.57];P<0.001). Those who received steroids had a higher chance of developing post-COVID-19 symptoms (1.86 [1.53-2.26];P<0.001). Time from recovery had no significant effect on the development of symptoms. On multivariate analysis, including hospitalization, steroid therapy and oxygen therapy, only those patients who received steroid therapy had higher odds of developing post-COVID-19 symptoms (1.86 [1.53-2.26];P<0.001).

## Discussion

The survey demonstrated that a significant proportion of patients recovered from COVID-19 suffer from post-COVID-19 symptoms. There are few reports of post-COVID-19 syndrome ^1,2^. Most studies reported fatigue and persistent respiratory symptoms ^2^. The reasons for developing post-COVID-19 symptoms are multiple: persistent chronic inflammation in the convalescent phase, sequelae of critical illness, organ damage (lung injury resulting in pulmonary fibrosis), and non-specific effects from the hospitalization and social isolation (psychological trauma, nutritional anemia, muscle wasting) ^2,3^. The limitation of our survey is we did not capture the data on age and comorbidities of the patients who reported symptoms. However, to our knowledge, this is the first Indian study to report a high incidence of post-COVID-19 symptoms among COVID-19 recovered individuals. Both hospitalized and non-hospitalized patients suffer from post-COVID-19 symptoms. Furthermore, the injudicious use of steroids led to a significant increase in the risk of post-COVID-19 symptoms.

## Conclusions

Post-COVID-19 symptoms are common in patients who recovered from COVID-19. These symptoms are more often noted in patients who received steroid therapy.

Post-COVID-19 symptomatology is present in a significant number of patients and requires to be addressed seriously.

## Data Availability

Data available on a reasonable request to the corresponding author.
https://forms.office.com/r/MGWKdJUpMk

## Acknowledgements

None.

---

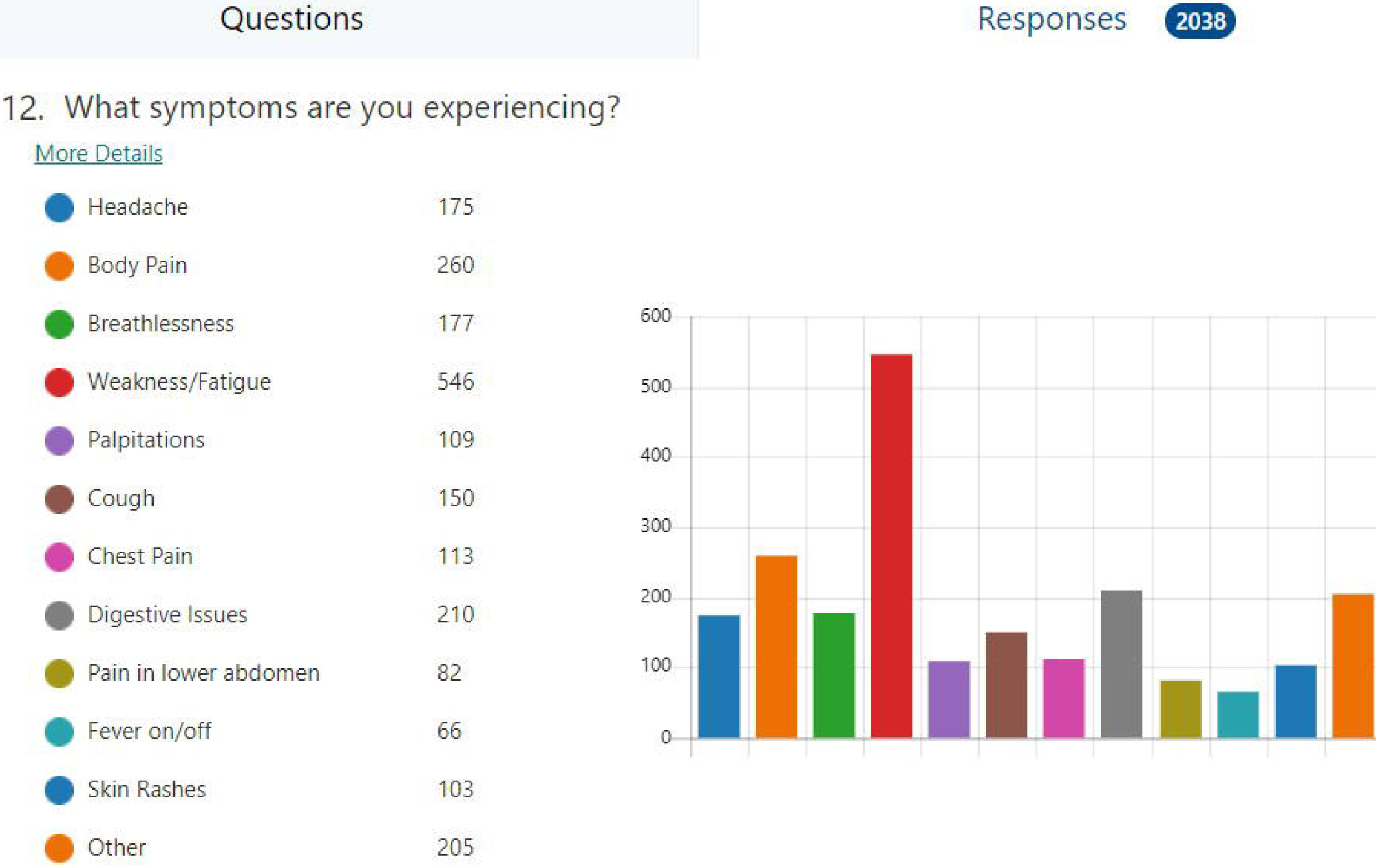

## References

1. Halpin SJ, McIvor C, Whyatt G, Adams A, Harvey O, McLean L, et al. Postdischarge symptoms and rehabilitation needs in survivors of COVID-19 infection: A cross-sectional evaluation. J Med Virol. 2021 Feb;93(2):1013–1022. doi: 10.1002/jmv.26368. Epub 2020 Aug 17. PMID: 32729939.

2. Yong SJ. Long COVID or post-COVID-19 syndrome: putative pathophysiology, risk factors, and treatments. Infect Dis (Lond). 2021 May 22:1–18. doi: 10.1080/23744235.2021.1924397. Epub ahead of print. PMID: 34024217; PMCID: PMC8146298.

3. Garg P, Arora U, Kumar A, Wig N. The “post-COVID” syndrome: How deep is the damage? J Med Virol. 2021 Feb;93(2):673–674. doi: 10.1002/jmv.26465. Epub 2020 Sep 29. PMID: 32852801; PMCID: PMC7461449.

4. Raveendran AV, Jayadevan R, Sashidharan S. Long COVID: An overview. Diabetes Metab Syndr. 2021 May-Jun;15(3):869–875. doi: 10.1016/j.dsx.2021.04.007. Epub 2021 Apr 20. PMID: 33892403; PMCID: PMC8056514.

