## Supplementary Material for "Post-COVID-19 symptoms are not uncommon among recovered patients-A cross-sectional online survey among the Indian population"

**Supplementary figure 1:** Respondents from the online survey.

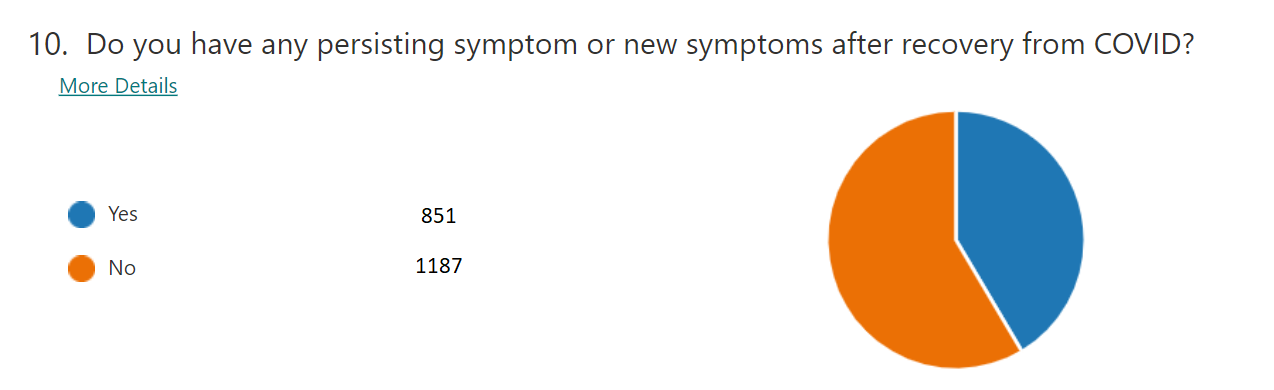

**Appendix: Questionnaire**

1. Name (optional)
2. Mobile number (optional)
3. City
4. How many days since you got recovered from COVID?
5. Did you require hospitalization because of COVID?
6. Did you require Oxygen support?
7. How many days you were hospitalized?
8. Did you receive Steroids?
9. How long did you continue your steroids regimen?
10. Do you have any persisting symptom or new symptoms after recovery from COVID?
11. Do you feel satisfied your post-COVID symptoms are being taken seriously?
12. What symptoms are you experiencing?
13. Did you get hospitalized because of Post-COVID complications?

Link to the survey:  <https://forms.office.com/r/MGWKdJUpMk>
